# Specific morphology of coronary artery aneurysms in mainly Caucasian Kawasaki Disease patients – Initial data from the Cardiac Catheterization in Kawasaki Disease registry

**DOI:** 10.1101/2023.10.19.23297286

**Authors:** Julia Weisser, Leonie Arnold, Wolfgang Wällisch, Daniel Quandt, Bernd Opgen-Rhein, Frank-Thomas Riede, Florentine Gräfe, Jörg Michel, Raoul Arnold, Heike Schneider, Daniel Tanase, Ulrike Herberg, Christoph Happel, Mali Tietje, Gleb Tarusinov, Jochen Grohmann, Johanna Hummel, André Rudolph, Nikolaus Haas, André Jakob

## Abstract

**Aims and Background:** Patients with a history of Kawasaki disease (KD), especially those with diagnosed coronary artery involvement, are known to require long-term cardiac care. However, specific evidence-based recommendations on long-term medical strategies are missing. Cardiac catheterization (CC) is still considered the gold-standard for diagnosing detailed coronary pathology. Therefore, and to better understand coronary artery pathology development in the long-term, we conducted a survey to document and evaluate CC data in a European population. Here we describe initial data on the first catheter examination these patients underwent.

**Method:** We administered a standardized questionnaire to retrospectively analyze CC data from KD children from the year 2010 until April 2023. This register covers basic acute phase clinical data and, more importantly, detailed information on morphology, distribution and the development of coronary artery pathologies. Data on participating departments of pediatric cardiology, mainly from Germany, were evaluated, with this study focusing on investigating each patient’s first CC exclusively.

**Results:** We analyzed a total of 164, mainly Caucasian, patients (65% male) across 14 pediatric cardiology departments. A relevant number of patients had no coronary artery aneurysm at the CC, facing the fact that distal CAAs were almost exclusively detected alongside proximal CAAs. Patients with multiple CAAs revealed a significant positive correlation between the number of CAAs and their dimensions, in diameter, as in length. Location of the CAA within the coronary artery, age at KD’s onset or gender did not significantly influence CAA diameters, but CAAs were longer in older children and in males.

**Conclusion:** The fact of distal CAAs being only present together with proximal ones, will hopefully reduce diagnostic CCs in KD patients without echocardiographically detected proximal CAAs. Furthermore this study gives valuable insights into dimensional specifics of CAAs in KD patients. As an ongoing register, future analyses will further explore long-term outcomes and performed treatments, helping to refine clinical long-term strategies for KD patients.

**Clinical perspective:**

- In Caucasian Kawasaki disease (KD) patients, distal coronary artery aneurysms (CAA) are usually diagnosed in association with proximal CAAs. Additional imaging such as conventional coronary artery angiography may therefore be unnecessary, if no aneurysms are detectable echocardiographically.
- In this context, potentially unnecessary CCs hopefully will reduce in future.
- Not only the CAA diameters, but also CAA length and CAA count may influence cardiac related morbidity and should be considered in long-term follow-up care.
- The age at the acute phase of KD, such as gender and location of CAA within the coronary arteries seems to not affect CAA dimensions.

**Clinical Trial Registration:** Cardiac Catheterization in Kawasaki Disease – data from the central European registry from 2010 – today

DRKS-ID: DRKS00031022, Date of Registration: 16.01.2023

## Introduction

The incidence of cardiovascular complications associated with Kawasaki Disease (KD) has significantly dropped^2, 3^ with improved early diagnosis and standardized therapeutic strategies, particularly the use of intravenous immunoglobulin (IVIG) and steroid treatment during the disease’s acute phase (AP). Cardiac involvement during the AP of KD was recently diagnosed in under 10%, with about a third of patients suffering from long-term cardiovascular sequelae. Though coronary artery dilatations/aneurysms (CAAs) are the main manifestations, valvular lesions, vascular stenosis and, rarely, myocardial infarction are also being reported^2, 4^.

Most CAAs caused by KD develop during its AP. Newly dilated coronary arterial lesions are rarely detected in KD’s late period. Various molecular etiopathogeneses seem to apply depending on the CAAs diameter, having an impact on long-term vascular sequelae. KD CAAs, especially those with large diameters, are primarily attributed to necrotizing arteritis characterized by severe inflammation and damage to the arterial walls. Due to chronic vasculitis, i.e. smaller aneurysms can also form in KD’s later stages and rarely even redevelop out of previously regressed CAAs^5^. Aneurysms with diameters considered to be small or medium are known to downsize, whereas giant CAAs do not tend to regress^6-9^.

Patients with a KD history, especially those with diagnosed coronary artery involvement, require long-term cardiac care. CC remains the gold standard for thorough coronary artery evaluation in adults; but for children with a history of KD, there is no universal consensus regarding CC’s diagnostic necessity. Existing KD-specific guidelines, with the American Heart Associatiońs (AHA) and Japanese Circulation Societýs (JCS) being the most consulted, recommend different approaches. The AHA’s guidelines, apparently taking a more restrictive approach on CC, suggest that it is optional for children with large coronary artery aneurysms (≥8mm/Z-Score ≥10) during the first year after the disease’s AP. According to the AHA, CC can be done as follow-up procedure to assess the progression or resolution of aneurysms over time for long-term monitoring of known persisting CAA. On the other hand, the JCS recommends that all patients diagnosed with a CAA exceeding 6 mm undergo at least one CC during the early reconvalescent phase of their disease. This suggests a more proactive approach to CC by recommending the procedure for a broader range of aneurysm sizes. The JCS also advises one follow-up CC when no further dilatation is echocardiographically apparent, most likely to confirm the former coronary pathology’s stability. German guidelines tend to follow the AHA’s risk stratification; there is international consensus that during KD’s acute inflammatory stage, interventional CC should only be done when there is evidence of myocardial ischaemia^2, 9-12^.

We have little evidence of long-term outcomes after KD, particularly of a predominantly Caucasian population. To address this gap, we initiated a register for CCs performed on patients with a history of KD in central Europe. This study was initiated by the working-group of interventional pediatric cardiologists endorsed by the German Society of Pediatric Cardiology and Congenital Heart Defects including participants from Austria and Switzerland. This register is open to any center to participate. A standardized questionnaire focused on detailed characterization of diagnostic CC outcomes, on interventions and the pharmacological strategies these KD patients experienced. In this present study we analyzed data on each reported KD patient’s first CC, focusing on a specific characterization of existing coronary artery pathologies.

## Material and Methods

This multicentric register study functions as a retrospective surveillance study for CCs done on children with a history of KD. The participating centers are requested to report data on all CC procedures conducted from 2010 onwards.

As of now, we have acquired data from 14 departments of pediatric cardiology, with 12 thereof in Germany, one from Switzerland and another from Sweden.

A standardized questionnaire addresses patients’ anthropometry as the main AP clinical characteristics, i.e., whether the CAA was already present during KD’s acute stage and whether IVIG had been administered. The questionnaire’s principal focus was on a detailed description of detected coronary artery pathologies: the number of CAAs present, including their specific location and each aneurysm’s diameter and length. As the AHÁs coronary segment classification^1^ was applied to assess the CAA’s location, pathologies were allocated not only to one of the coronary arteries’ main branches, but to one of 12 specific segments. This distinction enabled both more specific CAA location assessment and differentiation between proximal and distal CAAs. Striving for an age-and-body size-independent coronary diameter evaluation, the CAAs’ diameters were transferred to a coronary artery Z-Score via calculation proposed by Dallaire et al.^13^, acknowledging that these Z-Scores were originally applied for echocardiographic evaluation and primarily proximal coronary artery segments only. The aneurysm’s length was determined by its absolute extent. The data in this study focusses on the patients’ initial CC procedure.

### Statistical Analysis

The data’s distribution was tested using QQ plots and Shapiro-Wilk tests. Data are presented as median and interquartile range (IQR) or median and range. Categorical variables were reported as absolute numbers along with their respective percentages. To analyze the aneurysms’ extent, their maximum Z-Score was calculated also including their absolute diameters and length. We relied on the largest CAA (maximum diameter/length) or mean of all CAAs present (mean diameter/length) for our analysis of patients with multiple CAAs. The number of CAAs per patient was determined by adding together all CAAs regardless of their location. We ran a Kendall rank correlation to investigate the relationship between the CAA number/s per patient and maximum Z-Score or length. The coherence of the CAAs’ diameter and length, absolute dimensions and body surface area (BSA) adapted measurements, with the patients’ sex and their age at AP was analyzed by Kendall Tau, the Wilcoxon rank sum test and linear regression adjusted for BSA. Models for CAA dimensions and age at AP were not corrected for BSA due to a strong correlation between BSA and age at AP.

CAAs’ distribution per coronary artery branch was presented with their absolute count accompanied by their affiliated diameter and length. Z-Scores were presented as boxplots, one specifically assigned for each segment. For patients diagnosed with more than one CAA in a single segment, the larger one served for these size assessments. The Chi-square test was done to compare the occurrence of CAAs in proximal versus distal segments. Segments 1, 5, 6 and 11 were defined as proximal, CAAs affecting segments 2, 3, 4, 7, 8, 9, 10 and 12 were considered to be distally located. All data was analyzed in R 4.2.2 (R Core Team, 2022 Vienna, Austria).

## Results

### 1. General population

Our study includes a total of 164 patients, most of whom are male (65%). Median age at the time of catheterization was 3 years and 7 months; the time interval between the disease’s AP and patient’s first CC varied widely, ranging from month 0 to nearly 21 years. Median age at AP was 20 months, and 37.2% of the patients had been diagnosed with incomplete KD. Details on our patient cohort are in Table 1.

**Table 1:** General population data.

| **Patients' characteristics** |  |
| --- | --- |
| Number/sex of patients: n(m/f/missing) | 164 (107/38/19) |
| Diagnosis: Complete/incomplete KD/missing | 85/61/18 |
| Age at AP: Median (range) | 20 (0-243) months |
| Age at CC: Median (range) | 43 (0-312) months |
| Δ Time AP – 1. CC: Median (range) | 8 (0-251) months |

Specific questions addressed the existence of CAAs already present during the AP of KD. Put into relation to the number of CAAs present at time of AP, Figure 1 depicts CAAs detected at CC – revealing that 108 out of 138 (78,3%) had verified CAA early in the course of KD, but of all 164 patients, 77 (46.9%) had at least one CAA at their first CC. Among the patients initially diagnosed with CAA, approximately 48.1% (52 out of 108) no longer revealed coronary artery involvement via CC, indicating a relevant CAA regression rate. Only six patients had CAAs primarily detected during the CC, with five of them undergoing their initial CC within 4 months after the AP. An RCA aneurysm was detected at the CC performed 39 months after the AP of one patient only.

**Figure 1.**
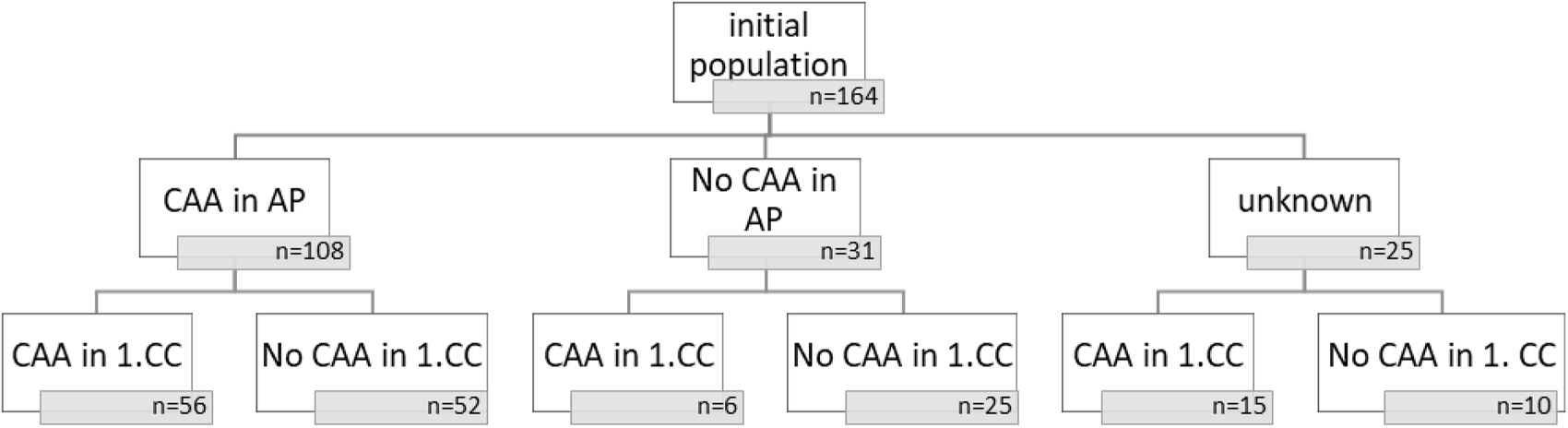
Flow chart – CAA in AP/ 1. CC; CAA= Coronary artery aneurysm, CC= Cardiac catheterization, AP= Acute Phase

### 2. CAA(s) – numbers and allocation

Further analysis focused on detailed descriptions of the CAAs detected in 77 patients. Regarding the number of CAAs per patient, 57 out of 77 (74.0 %) had multiple CAAs. Two simultaneous CAAs were diagnosed in 22 patients, three in 20 patients. 10 patients were diagnosed with more than four simultaneous CAA, with eight the highest number of CAAs in a single patient.

With many patients having numerous CAAs, our study cohort comprises a total of 203 CAAs. With 87 CAAs located in the RCA and 116 CAAs in the left coronary artery, the distribution of these aneurysms is roughly equal between the two sides. To more precisely allocate the CAAs, we assigned them to one specific coronary artery segment applying the AHA’s classification^1^. CAAs continuously extending over more than one segment qualified as multiple segments. The exact number of aneurysms identified in each segment is found in Figure 2. The proximal RCA (segment 1) is the one most frequently affected by CAAs, followed by the proximal part of the left anterior descending coronary artery (LAD) (segment 6). Distal coronary artery segments seldom reveal CAAs according to our register’s data.

Not only do distal segments not tend to develop CAAs, there is distal coronary artery involvement almost exclusively when a CAA is also detected in one of the respective proximal segments (see Table 2). We further analysed the presence of distal CAAs in relation to coexisting proximal CAAs in any of the main coronary arteries, identifying only one patient who seemed to have developed an isolated distal RCA-CAA (location: segment 3; dimensions: diameter 4.4 mm/Z-Score 5.9 x length 4.4mm). However, this one was a “non-detected KD” patient suffering from acute coronary ischemia caused by a complete thrombotic LAD-occlusion, he was assumed to have also had an underlying corresponding proximal LAD aneurysm. None of our register patients presented an isolated distal CAA.

**Table 2:** Occurrence of proximal and distal CAAs. Isolated distal CAAs vs. distal and concurring proximal CAAs in the same coronary artery (p-values according to Fisher’s exact test)

| **Proximal vs. distal CAA occurrence** |  |  |  |  |
| --- | --- | --- | --- | --- |
|  | Only prox. | prox. and distal | Only distal | Prox. And distal vs. only distal |
| RCA | 38 | 18 | 2 | p-value<0.001 |
| LAD | 39 | 10 | 0 | p-value<0.001 |
| LCX | 19 | 1 | 2 | p-value<0.001 |

### CAA(s) – sizes and dimensional distribution

Regarding CAA dimensions, we documented a significant range of CAA sizes in diameter and length. The largest CAA was located in the RCA, measuring 33.0 mm (Z-Score 78.3) in diameter and 85.0 mm in length. Each coronary artery’s specific CAA dimensions are listed in Table 3. Besides the largest aneurysm present in the RCA, we found no significant difference in CAA diameters and lengths among all the main coronary arteries.

**Table 3:** CAA distribution with dimensional measurements.

| **CAAs' characteristics per coronary artery main branch** |  |  |  |  |
| --- | --- | --- | --- | --- |
|  | RCA | LCA | LAD | LCX |
| Total number (n) | 87 | 29 | 61 | 26 |
| -with size specification (n) | 83 | 25 | 56 | 21 |
| Diameter (mm): Median (range) | 6.0 (1.9-33.0) | 5.8 (2.5-25.0) | 5.5 (2.2-22.0) | 4.0 (2.3-11.0) |
| Z-Score: Median (range) | 9.7 (2.5-78.3) | 8.6 (3.4-65.8) | 8.4 (2.6-46.9) | 5.7 (2.8-27.1) |
| Length: Median (range) | 9.0 (2.0-85.0) | 8.5 (3.0-30.0) | 8.0 (1.3-85.0) | 6.0 (2.7-30.0) |

Although proximal segments develop CAAs more frequently, the aneurysm’s size does not seem to be influenced by its distance from the coronary artery’s origin, meaning that proximal CAAs are on average not significantly larger than more distally located ones. However, giant-sized CAAs are indeed primarily detected in proximal coronary artery segments (Figure 3).

**Figure 3.**
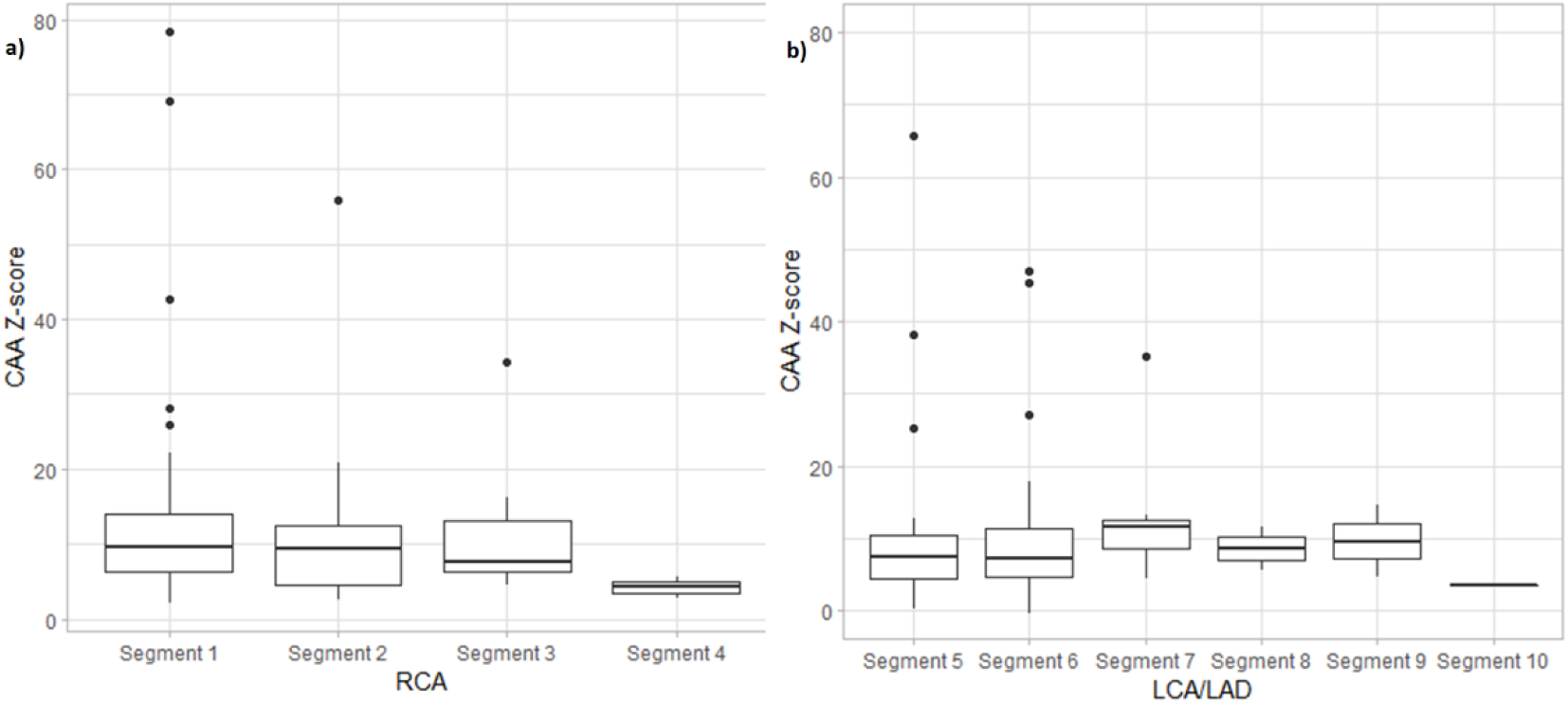
Boxplots of CAA diameter per segment of the a) RCA: Median/IQR: Seg 1 10.07/6.90, Seg 2 9.10/7.60, Seg3 7.51/6.90, Seg 4 4.21/1.40, b) LCA/LAD: Median/IQR: Seg 5 7.31/5.97 Seg 6 7.18/6.40, Seg 7 11.49/3.94, Seg 8 8.55/3.06, Seg 9 9.61/4.99, Seg 10 3.46/0.00; CAA= Coronary Artery Aneurysm, RCA= Right coronary artery, LCA= Left coronary artery, LAD= Left anterior descending coronary artery; IQR= Interquartile range; Seg= Segment

We also analyzed potential CAA size-influencing factors. The number of CAAs diagnosed in a single patient indicates a significant positive correlation with both CAA diameter (R=0.45, p=0,0031) and its length (R=0.34, p = 0.00045). The more aneurysms a patient has, the larger they are (see Figure 4).

Moreover the influence of sex and age during the AP of KD on CAA development was investigated. As expected, older patients present larger CAAs than younger patients in absolute-dimension terms, demonstrated by a moderately significant correlation between age and CAA dimensions. This correlation disappears when accounting for a patient’s BSA via coronary artery Z-Scores. We also examined the influence of gender and age on the diameter of each patient’s largest (or only) CAA (max. diameter/length) and their patient-specific means (mean diameter/length). In terms of the aneurysms’ length, there is no adjustment for BSA. However, our register’s data showed that the older the patients are, the longer their CAAs are as well. Furthermore, boys seem to develop significant longer CAAs than girls. The total number of CAAs on the other hand, does not seem to be influenced by either sex or age at disease onset (see Table 4).

**Table 4.**
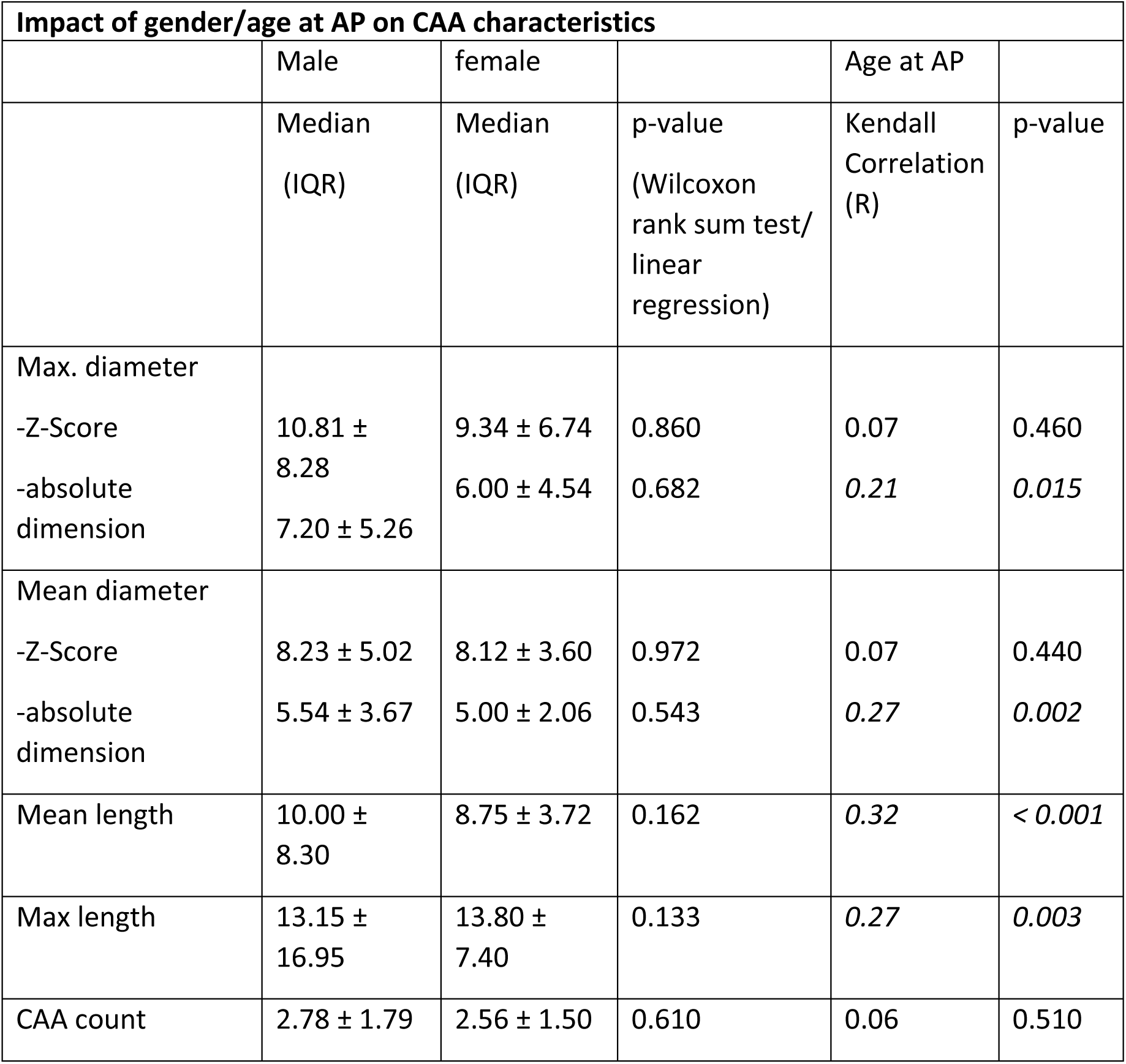
Median diameter/length and numbers of all CAAs per patient comparing boys and girls indicating a significant difference in CAA length and Kendall’s correlation coefficient (R) with patient age at AP indicating no significant correlation between body size adapted Z-scores/CAA count and the age. CAA= coronary artery aneurysm, IQR= Interquartile range.

## Discussion

This study describes a total of 164 patients from several pediatric cardiology departments. With two university hospitals outside Germany participating, this register study represents KD in mainly Caucasian patients. Our data analysis reveals several key findings. This study demonstrates a considerable regression rate in the numbers of CAAs when comparing CAA numbers from the AP to those in the first CC, confirming established evidence from older Japanese CC data^7, 8^. The development rate of new CAAs is low throughout their later stages – 3% according to our data. As our data analysis concentrated mainly on elaborating upon CAA morphology based on CC measurements, only patients with available data from specific CAA(s), such as size and location, were included in further analyses. Studying the numbers of CAAs locally, we identified aneurysms distributed about equally to both coronary arteries, with the proximal RCA segment revealing most CAAs. Distal CAAs were diagnosed almost exclusively when there was an additional coronary artery pathology in a proximal segment – in fact, this cohort presented no isolated distal CAA. The rarity of distal aneurysms seems to be generally accepted, as it is also reflected in the AHA KD guidelines^9^. However, there is no concrete reference, and little research has been done on this topic to support this finding. Most data relies on CT imaging, confirming distal-only CAAs as rare, as did a recent study evaluating CT coronary angiography in Indian children with KD^14^. Their study included 23/176 patients diagnosed with distal CAA(s), four of them had isolated distal coronary artery pathologies. A coexisting proximal CAA was detected in 19 other patients, or proximal CAAs that contiguously affected also distal segments. Despite these findings, there is a paucity of reliable data on the occurrence of distal KD-linked coronary artery abnormalities, with the great majority of current data from findings from studies on Asian children. The significant difference in KD incidence among ethnicities, i.e. 254/100.000 in Japan and 6-7/100.000 in Germany highlighting the importance of ethnic-specific data, especially for KD children of predominantly Caucasian origin^9, 15, 16^.

A considerable number of our patients suffered from multiple simultaneous CAAs, with their number correlating positively with their CAÁs diameters and lengths. Boys and older children reveal significantly longer CAAs (noting that these specific parameters were not BSA-adjusted). The aneurysm’s length, especially its pathological impact on cardiac morbidity, has not been adequately assessed to date. The number and diameters of CAAs do not seem to be influenced by either age during the disease’s AP or the patient’s gender. Moreover, our data indicates that CAA size seems to be independent of its location. CAA dimensions did not differ in relation to the segment in which they had developed. This may indicate that although distal CAAs are rarer, they can still develop large dimensions and thus have a direct impact on KD patients’ long-term clinical outcomes.

The understanding of CAA pathogenesis in KD remains limited. Initially, mouse model data indicated that certain factors, including IL-ß117 and Single Nucleotide Polymorphisms (SNPs) in various genes, such as inositol 1,4,5-trisphosphate 3-kinase C (ITPKC) and the TIFAB gene, play a role in predisposing individuals to KD and increasing the risk of CAA formation^17-20^. Additionally, the expression of CD40 Ligand on CD4+ T-cells and platelets, as well as an imbalance in the levels of matrix metalloproteinases (MMPs) and tissue inhibitors of MMPs, may also influence the extent of coronary artery involvement in KD^21^.

While some data on the acute formation of CAAs exist, our molecular knowledge of long-term CAA development is notably lacking. A comprehensive understanding of the morphology of CAAs in the long term is essential not only for optimizing long-term clinical management but also for shedding light on the molecular pathomechanisms underlying CAA development.

Monitoring and correctly assessing the severity of CAAs in KD patients, echocardiography is the preferred noninvasive diagnostic tool. It is performed at the disease’s onset and should be repeated according to the patient’s clinical presentation and follow-up evaluation later on. While it effectively reveals coronary artery involvement and usually enables assessment of the disease’s severity, its sensitivity and specificity in determining distal coronary artery involvement as for detecting coronary artery stenosis and thrombosis are limited^22^ ^23^. CC is generally still considered the “gold standard” for thorough evaluation of coronary arteries in KD. However, CTCA has proven to be of similar diagnostic efficacy as CC. In fact, CTCA may even be more precise, particularly for visualization of distal coronary arteries and intramural anomalies^24-28^. Radiation exposure is considered to be similar in both procedures^28^. MRCA, while radiation-free, has limited capacity to visualize distal segments, but it may be beneficial for patients with severe intramural calcifications, providing simultaneous information on cardiac function ^28^.

This register study primarily aimed for a detailed (not favorizing) review of CC findings in European patients diagnosed with KD specifically. CC can help to determine the severity of KD’s cardiac involvement, but alternative angiographic methods (also as substitute strategies), should always be considered keeping CC’s strengths and limitations in mind. As a general rule, the application of CC use should be clearly indicated especially as proximal CAAs are sufficiently diagnosed echocardiographically. The number of possibly non-essential CCs seems to have already dropped over the years. Comparing this study’s data to Japanese cardiac catheterization data from of the 1970s^8^ and -80s^13^ (when echocardiography was not universally available or precise enough), the rate of CCs revealing only the absence of coronary artery damage has more than halved. Stricter indications and better alternative evaluation methods can be considered potentially causative factors behind this trend. Although we still lack clear guidelines (for central European patients specifically), the number of CCs done in children presenting no coronary artery involvement (88 of 164 = 53,7%) remains fairly high. Considering CCs done mainly between 2010 and 2022, note that the mean rate of CCs without any CAA detection per year varied from 62,5% in 2011 and 25,0% in 2022, indicating a reduction in potentially unnecessary CCs. We hope with the data provided, future analysis hopefully will demonstrate a further and significant drop of this number.

Data of this study was acquired retrospectively by standardized questionnaires, data quality therefor depends on the reporting physicians themselves and could not be uniformly verified. Furthermore few aspects such as the CC’s indication were not evaluable, mostly applying to open question types. Aiming for a high total of analyzed CCs (and that our registry study’s primary aim was to address CC findings), we were only able to sketch other disease aspects such as AP data and treatment strategies. We could not compare angiographic CAA measurements to their dimensions during the AP, as initial echocardiographic data was not universally available. Nevertheless our register was established to primarily collect specific, mostly CC related data on mainly Caucasian children suffering from KD, giving valuable insights to specific CAA data, to optionally influence local adaptations of follow-up clinical strategies.

## Data Availability

Data, consistent with the declareation of Helsinki and German data protection law, are available upon request.

## Non-standard Abbreviations and Acronyms

KD: Kawasaki Disease
CC: Cardiac Catheterization
CAA: Coronary Artery Aneurysm
AP: Acute Phase
IVIG: Intravenous Immunoglobulin
AHA: American Heart Association
JCS: Japanese Circulation Society
IQR: Interquartile Range
BSA: Body Surface Area
RCA: Right Coronary Artery
LCA: Left Coronary Artery
LAD: Left Anterior Descending Coronary Artery
LCX: Left Circumflex Coronary Artery
SNP: Single Nucleotide Polymorphism
ITPKC: Inositol 1,4,5-trisphosphate 3-kinase C
MMP: Matrix Metalloproteinases

## Acknowledgments

We thank all participating centers for providing valuable data and detailed information on cardiac catheterizations. We also thank the German Society of Pediatric Cardiology for supporting this registry.

